# Forecasting Acute Kidney Injury and Resource Utilization in ICU patients using longitudinal, multimodal models

**DOI:** 10.1101/2024.03.14.24304230

**Authors:** Yukun Tan, Merve Dede, Vakul Mohanty, Jinzhuang Dou, Holly Hill, Elmer Bernstam, Ken Chen

## Abstract

**Background:** Advances in artificial intelligence (AI) have realized the potential of revolutionizing healthcare, such as predicting disease progression via longitudinal inspection of Electronic Health Records (EHRs) and lab tests from patients admitted to Intensive Care Units (ICU). Although substantial literature exists addressing broad subjects, including the prediction of mortality, length-of-stay, and readmission, studies focusing on forecasting Acute Kidney Injury (AKI), specifically dialysis anticipation like Continuous Renal Replacement Therapy (CRRT) are scarce. The technicality of how to implement AI remains elusive.

**Objective:** This study aims to elucidate the important factors and methods that are required to develop effective predictive models of AKI and CRRT for patients admitted to ICU, using EHRs in the Medical Information Mart for Intensive Care (MIMIC) database.

**Methods:** We conducted a comprehensive comparative analysis of established predictive models, considering both time-series measurements and clinical notes from MIMIC-IV databases. Subsequently, we proposed a novel multi-modal model which integrates embeddings of top-performing unimodal models, including Long Short-Term Memory (LSTM) and BioMedBERT, and leverages both unstructured clinical notes and structured time series measurements derived from EHRs to enable the early prediction of AKI and CRRT.

**Results:** Our multimodal model achieved a lead time of at least 12 hours ahead of clinical manifestation, with an Area Under the Receiver Operating Characteristic Curve (AUROC) of 0.888 for AKI and 0.997 for CRRT, as well as an Area Under the Precision Recall Curve (AUPRC) of 0.727 for AKI and 0.840 for CRRT, respectively, which significantly outperformed the baseline models. Additionally, we performed a SHapley Additive exPlanation (SHAP) analysis using the expected gradients algorithm, which highlighted important, previously underappreciated predictive features for AKI and CRRT.

**Conclusion:** Our study revealed the importance and the technicality of applying longitudinal, multimodal modeling to improve early prediction of AKI and CRRT, offering insights for timely interventions. The performance and interpretability of our model indicate its potential for further assessment towards clinical applications, to ultimately optimize AKI management and enhance patient outcomes.

## 1. Introduction

AKI, characterized by a substantial elevation in serum creatinine (SCr) or a pronounced decline in urine output^1^, represents a severe clinical complication marked by a sudden decline in kidney function. AKI is a profoundly serious condition and needs to be intercepted immediately to prevent lasting kidney damage. This condition is linked to elevated hospital costs and extended length of stay^2–4^, with a prevalence exceeding 50% among intensive care units (ICU) patients and accompanied by an alarming mortality rate, up to 50% within the ICU settings^5,6^. The International Society of Nephrology^7^ emphasizes the significance of early identification of individuals at increased risk of developing AKI for potentially better outcomes. Early detection allows for therapeutic intervention before the onset of complications, such as anuria, and associated issues like acidosis, hyperkalemia, volume overload, as well as long-term complications such as lung injury, sepsis, and chronic kidney disease.^8–11^. Beyond the immediate health implications, AKI significantly impacts healthcare resource allocation, particularly in the context of dialysis. Among available dialysis techniques, Continuous Renal Replacement Therapy (CRRT) is the most commonly used method for hemodynamically unstable patients, accounting for up to 75% of instances in ICUs^12^. However, the decision to initiate CRRT is influenced by multiple factors. While there are guidelines and recommendations, there is not standardized criteria. Instead, the decision often depends on specific clinical context and individual patient conditions, highlighting the complexity of managing AKI-induced resource utilization and underscoring the need for a comprehensive approach to enhance clinical decision-making and resource allocation strategies.

The widespread adoption of EHRs and the rapid advancements in computing offer new avenues for disease prediction and prevention. These advancements have been particularly transformative in developing integrated clinical decision support systems^13,14^. The volume of routine data collection during hospital stays, considering its high temporal resolution, often exceeds human cognitive processing capabilities^15^. In response to this data complexity, Artificial Intelligence (AI) emerges as a promising solution to effectively process such large-scale, high-dimensional datasets. This is particularly relevant in the context of AKI and CRRT prediction. AKI, characterized by its well-defined temporal progression and CRRT, are especially suited for EHR-based predictive modeling^16^, utilizing techniques such as logistic regression (LR), Extreme Gradient Boosting (XGBoost), Transformer and Long Short-Term Memory (LSTM). Meanwhile, in the realm of clinical records, researchers have actively sought to harness the capabilities of natural language processing (NLP) for medical prediction tasks. This pursuit has led to the exploration of various methodologies, including a convolutional document embedding method grounded in the unstructured textual content of clinical records^17^, an open set of biomedical word vectors/embeddings that integrates domain-specific biomedical knowledge^18,19^, and the latest Bidirectional Encoder Representations from Transformers (BERT) model^20^ which addresses the limitation of representing each word with only one vector^21^. Although several AKI or CRRT prediction models have been reported recently, notable limitations exist. First, these models have often been derived and validated within specific patient cohorts, such as those undergoing cardiac surgery or catheterization^22–25^, children^26^, elderly adults^27^, or the critically ill^28^. This specificity may limit their generalizability to broader patient populations. Second, a common short fall in these models is their reliance on static features or single-point measurements^29–32^. This approach does not fully capture the dynamic and rapidly evolving nature of patients’ health status, especially in the context of AKI development as well as in postoperative settings where physiological parameters can change quickly and unpredictably. The need for models that adapt to these rapid changes in a patient’s condition is crucial for accurate AKI and CRRT prediction. Third, many studies do not account for non-zero gap times in their predictive frameworks, leading to the risk of temporal data leakage of label information during model training^24,33,34^. For instance, utilizing fixed 6-hour input windows may inadvertently include data points just before the AKI diagnosis, potentially skewing the prediction by using information already known to the care team. Additionally, most existing models are unimodal, focusing either solely on laboratory measurements or clinical notes, thus lacking the comprehensive view provided by a multimodal approach. This limitation is particularly critical in scenarios, where data availability is constrained or highly imbalanced. Finally, it is noteworthy that a considerable portion of existing literature tends to limit its predictive analyses by relying on basic measurements, thus failing to optimize the full potential of the available datasets.

To address these gaps, we have developed a multi-modal model that integrates the embeddings of LSTM and BioMedBERT. This advanced model leverages a blend of both unstructured clinical notes and structured time series measurements, incorporating over 50 features enhanced by medical insights derived from EHRs. It enables the early prediction of both AKI and the anticipation of the need for CRRT by providing a minimum of 12-hour lead time before clinical manifestation. Recognizing the clinical need for actionable insights, our model is designed not just to predict AKI earlier and with greater accuracy but also to influence ICU interventions. By identifying patients at high risk for AKI/CRRT, our model can potentially inform clinical decisions to minimize renal risk, such as the judicious use and dosage of nephrotoxic drugs, careful consideration of imaging procedures involving ionizing contrast dye, avoiding hypotension, and meticulous management of hydration levels. Understanding a patient’s heightened risk of renal failure recalibrates the risk/benefit ratio of common ICU interventions, allowing clinicians to consider treatment strategies that reduce the likelihood of AKI and the need for CRRT, thereby potentially improving patient outcomes and resource utilization.

|  |  |
| --- | --- |
| Problem or Issue | Forecasting AKI and the need for CRRT in ICU patients. |
| What is Already Known | Existing AKI/CRRT prediction models are limited in: often tailored to specific patient groups like cardiac surgery patients, relying on static features or single-point measurements, ignoring non-zero gap times, and using unimodal frameworks. |
| What this Paper Adds | We developed a machine-learning, longitudinal, multi-modal model that leverages both clinical notes and time-series measurements. Our model enables early prediction of AKI and CRRT with at least 12 hours' lead time. Furthermore, our model identifies previously overlooked factors contributing to AKI and CRRT need. |

## 2. Materials and methods

### 2.1 Dataset description

The dataset utilized in this study was sourced from the MIMIC-IV database, spanning a period from 2008 to 2009 and including intensive care unit (ICU) patient stays at Beth Israel Deaconess Medical Center. The database comprises a comprehensive array of patient-related information, including demographics, laboratory test results, procedures, medications, clinical notes, imaging reports and mortality data, encompassing post-hospital discharge outcomes (Figure 1(a)). Within the MIMIC-IV database, patients are denoted as Subject_IDs, with each patient having one or more hospital admissions named Hadm_IDs. Within a single admission, a patient may undergo one or more ICU stays, referred to as Icustay_IDs. A clinical event is defined as an individual measurement, observation, or treatment.

**Figure 1.**
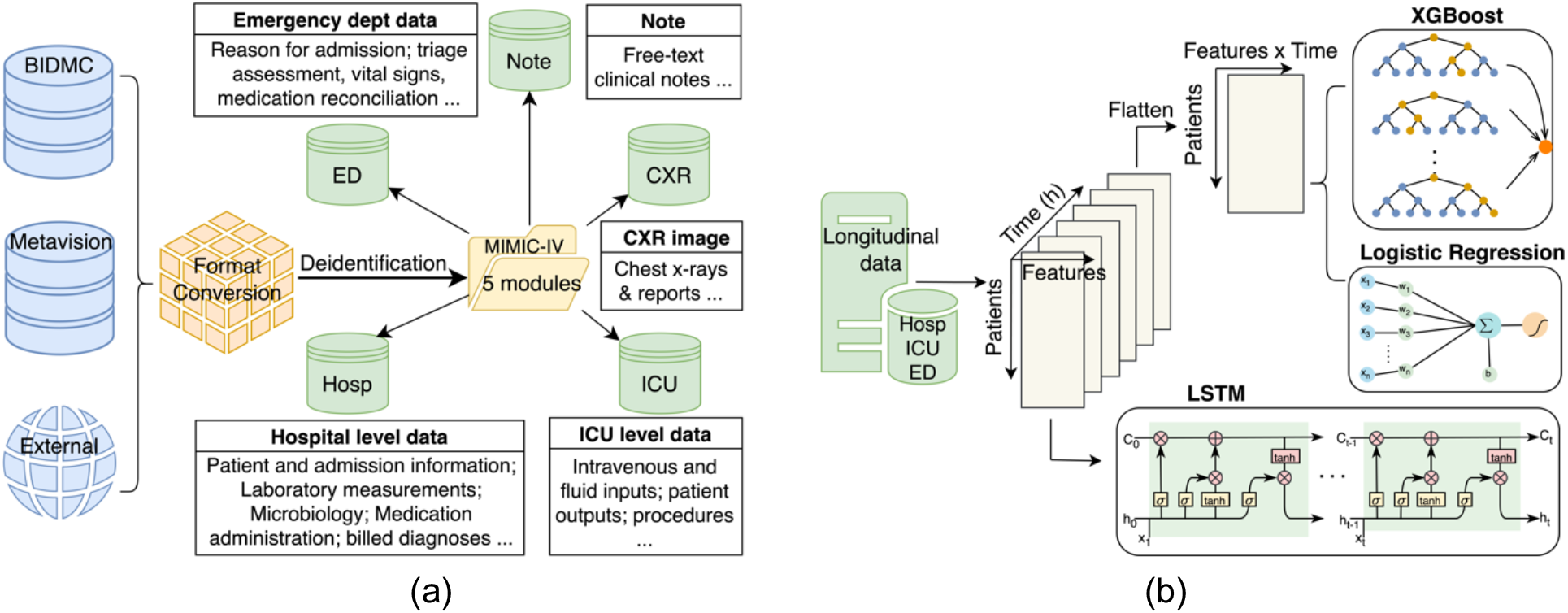
(a) MIMIC-IV built by merging the various data sources (the BIDMC data warehouse, the ICU information system, and external sources) into a single schema with five modules. (b) Comparative analysis of three model types (XGBoost, Logistic Regression, LSTM) for processing 6-hour time series lab measurement data.

### 2.2 Methodological framework

In this study, we adopt the classification system proposed by the Acute Kidney Injury Working Group of the Kidney Disease: Improving Global Outcomes (KDIGO) Foundation to standardize the definition of AKI. As per the KDIGO criteria, AKI is categorized into three stages^1,35,36^, with Stage 1 indicating the initial phase of AKI, often asymptomatic. Stages 2 and 3 signify increasing severity of renal dysfunction. Our research focuses on patients diagnosed with Stage 2 and Stage 3 AKI, given their clinical significance and the progressive nature of the condition at these stages. Furthermore, we extend our analysis to predict the need for CRRT.

The predictive model we propose is specifically designed to analyze time series laboratory measurements collected in the first 6 hours post-admission. Our goal is to forecast the probability of a patient being diagnosed with AKI or requiring CRRT within a 12-hour period after this initial 6-hour data collection window. This design ensures that our predictions apply to patients diagnosed at least 18 hours after admission, thereby reducing the risk of predicting AKI in individuals who might have been diagnosed prior to hospitalization.

In terms of clinical notes, our model selectively incorporates notes recorded at the time of patient admission. This includes information of the chief complaint, history of present illness, past medical history, initial physical examination findings, and initial clinical impressions. Importantly, we exclude discharge notes to focus on data relevant to the initial phase of hospitalization, therefore enhancing the model’s precision and clinical utility.

### 2.3 Time series embedding model

We designed the Time Series Embedding Model to acquire the temporal representation of patients utilizing 50 distinct features selected based on medical knowledge. Our investigation delved into three prominent classification methodologies: logistic regression (LR), Extreme Gradient Boosting (XGBoost), and Long Short-Term Memory (LSTM). These models are widely acknowledged and extensively employed in the biomedical domain, as depicted visually in Figure 1(b). While Transformer-based architectures^37–39^, have gained significant traction in recent years, their substantial data requirements, e.g., included several million samples in^38^, make them less suitable for our investigation.

#### 2.3.1 Logistic regression (LR)

LR is characterized as a linear classification model with limited complexity and moderate interpretability. Due to its inherent limitation in handling temporal data, we concatenated 6 one-hour measurements from patients into a single vector. In this model, we used a more elaborate version of the hand-engineered features^40,41^, incorporating six distinct sample statistic features for each variable within a given time series. These features encompass the minimum, maximum, mean, standard deviation, skew, and the number of measurements within each subsequence.

#### 2.3.2 Extreme Gradient Boosting (XGBoost)

XGBoost is a state-of-the-art machine learning algorithm utilizing gradient-boosting trees for predictive modeling^42^. Its notable advantages include high predictive accuracy, automatic modeling of non-linearities and high-order interactions, as well as robustness to multicollinearity. Similar to LR, we utilized XGBoost to process temporal data by concatenating 6-hour measurements into a single vector.

#### 2.3.3 Long Short-Term Memory (LSTM)

LSTM models^43^, a popular variant of recurrent neural networks (RNNs), exhibit the capacity to process sequences of arbitrary lengths in a non-linear and high-capacity manner. This enables the learning of sequential relations and long-term temporal dependencies within time series data. The crucial role in achieving this function lies in the gate and state of the network, with the forget gate determines whether to delete the previous state. The final hidden state of the LSTM serves as the time series embedding, utilized for subsequent predictions.

It is noteworthy that both LR and XGBoost generate predictions based on a flattened vector of the 6-hour time series data, while LSTM can directly process the time series input to make binary predictions. Our results have shown that the LSTM model outperforms the other two models — LR and XGBoost (Table 1). Consequently, we have made the informed decision to incorporate the LSTM model as a crucial component of our multimodal model.

**Table 1.**
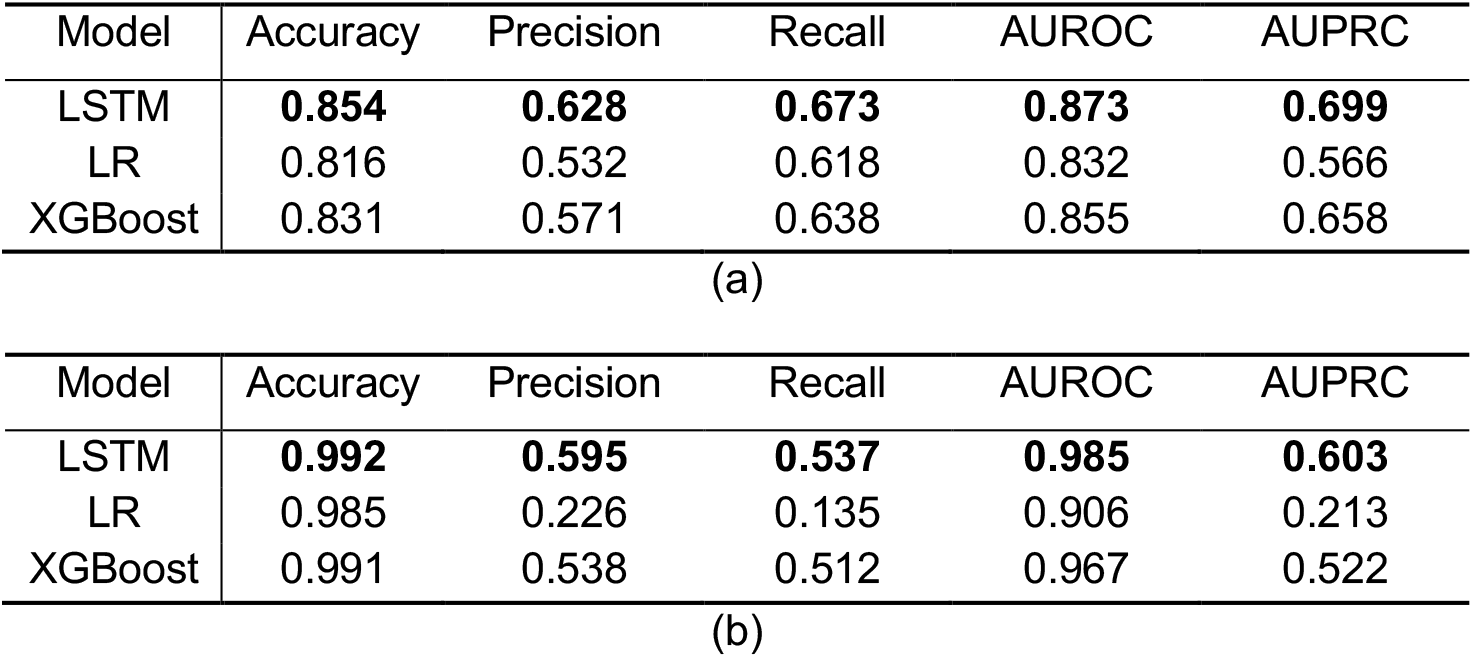
LSTM, LR, and XGBoost performance comparison for (a) AKI (b) prediction.

### 2.4 Clinical notes embedding model

Our study delves into the exploration of two established BERT-based models, namely BioMedBERT and ClinicalBERT, specifically tailored to enhance natural language processing within the biomedical and clinical literature domains. We enhance this capability through the fine-tuning process on specific tasks, thereby constructing a specialized BERT-based AKI/CRRT prediction model.

#### 2.4.1 BioMedBERT^44^

BioMedBERT, previously named PubMedBERT, specially pretrained from scratch using abstracts from PubMed and full-text articles from PubMed Central, excels in capturing the nuances of medical terminology, scientific jargon, and the vast array of topics within the biomedical field. It has demonstrated remarkable performance in tasks, such as document classification and information retrieval within the biomedical domain.

#### 2.4.2 ClinicalBERT^45^

ClinicalBERT focuses on clinical notes/EHRs, and other healthcare-related documents. By training a BERT model on clinical text, ClinicalBERT has been optimized to understand the specific language nuances present in patient records, medical reports, and uncovers relationships between medical concepts that match physician judgment.

#### 2.4.3 Probability calculation of solely BERT-based model

The computation of predictions for patients with an extensive volume of clinical notes entails a binning process, wherein predictions are aggregated for each subsequence. In the context of a patient whose notes are segmented into n subsequences, the BERT-based model generates a probability estimate for each respective subsequence. As per the methodology established by Huang^45^, the probability of AKI and CRRT for a given patient is expressed as:

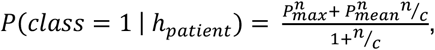

where *c* is the scaling factor to control over the influence of the number of subsequences *n*, and *h*_*patient*_ denotes the implicit representation encompassing all notes associated with a specific patient. The probabilities 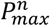 and 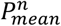 represent the maximum and mean probabilities of AKI/CRRT over *n* subsequences.

The underlying concept posits that certain subsequences may lack informative content regarding AKI/CRRT, while others are relevant. Therefore, computation of AKI/CRRT risk is selectively performed using subsequences that exhibit correlation with the target outcome, thereby minimizing the impact of less informative subsequences. However, it is worth noting that maximum probability may be caused by noise. In this context, we include the mean probability 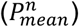 to have a trade-off between the mean and maximum probabilities. Finally, to adjust for patients with varying quantities of clinical notes, we introduce a scaling factor *n*/*c*, providing a larger weight for the mean prediction in cases where patients possess a higher volume of notes. Notably, empirical findings by Huang^45^ indicate that *c*=2 yields optimal performance based on validation data.

The comparison results from the experiment section reveal that BioMedBERT has a superior performance compared to ClinicalBERT. Notably, one of the key differentiators lies in the foundational pretraining approach employed for each model. BioMedBERT is pretrained from scratch, allowing it to capture intricate nuances and domain-specific intricacies. On the other hand, ClinicalBERT is built upon the BERT model, with its training grounded in general-domain language models. As a result, we decided to integrate the BioMedBERT model as an essential element within our multimodal framework.

### 2.5 Multimodal Learning

All those models above use only one data source to make predictions. Previous studies show that the combination of time series and clinical record is useful for the prediction^18,46,47^. Therefore, we used a representation of the clinical record in conjunction with the time series portion of the patient data and incorporated the fusion module to improve the performance of the prediction.

As depicted in Figure 2, our proposed model seamlessly integrates two distinct modalities. Specifically, we concatenate the embeddings of two top-performing unimodal models—LSTM for time-series measurements and BioMedBERT for clinical notes. The multimodal encoder harmonizes these embeddings, fusing the information from both modalities and projecting them into a shared space.

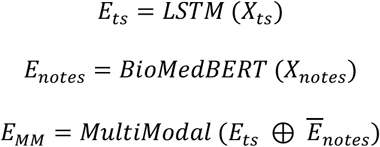

**Figure 2.**
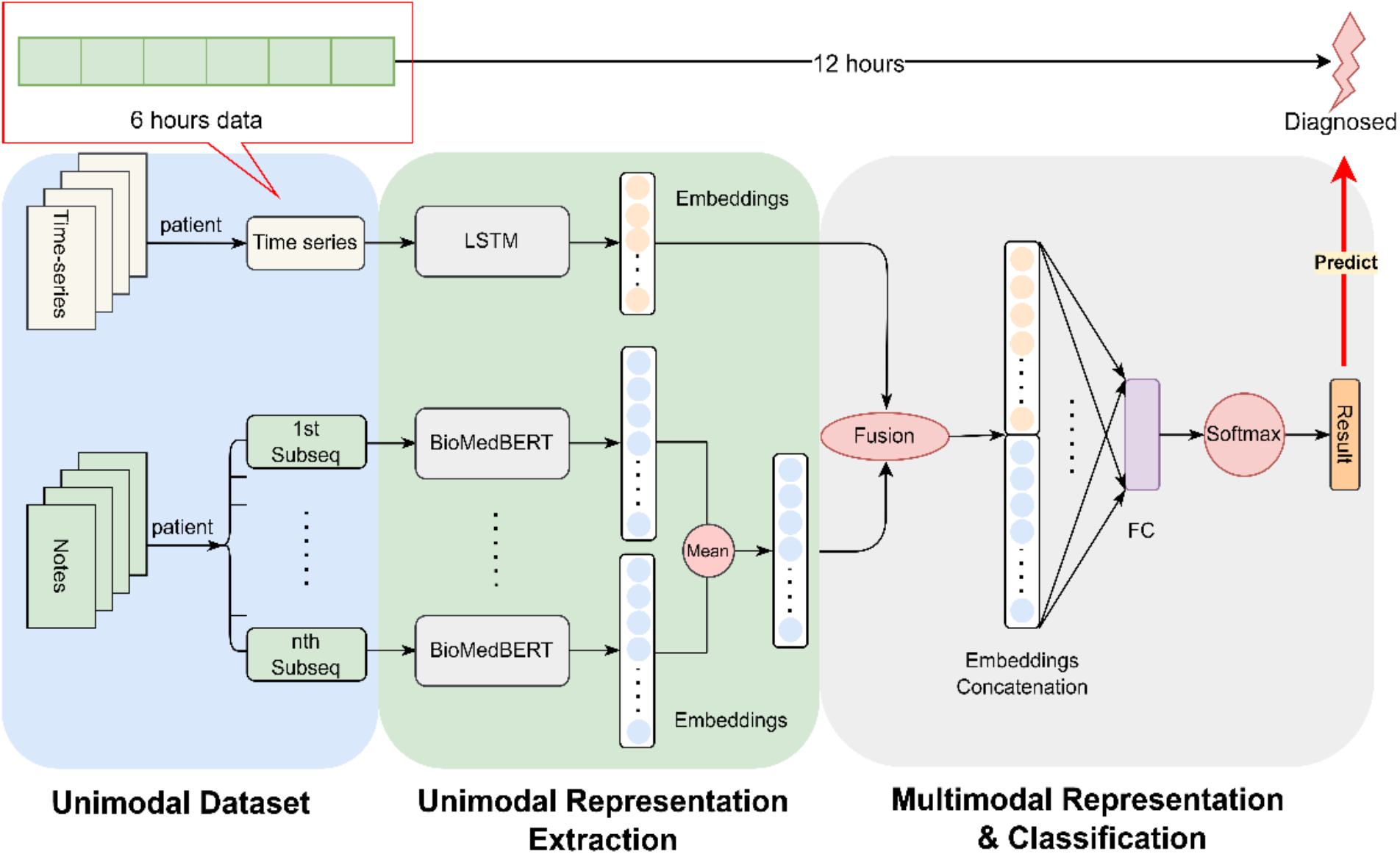
Schematic Representation of the Proposed Multimodal Model: The diagram illustrates our proposed framework, wherein we concatenate embeddings derived from 6 hours’ time-series measurements using LSTM and clinical notes processed by BioMedBERT. This combined representation is utilized for the prediction of AKI/CRRT, aiming to forecast clinical manifestation at least 12 hours in advance.

In this study, we utilize time series measurements denoted as *X*_*ts*_ ∈ *R*^2×4^, where *X*_*ts*_ represents the time series measurements with L length of measurements counted by hours, and *D* variables. In our case, L is set to 6, and D is 100, as following Harutyunyan’s setup^41^ that *X*_*ts*_ is composed of original time series variables and binary time series variables which indicates whether the corresponding variables are observed or imputed. The details regarding data curation are described in the experiment section.

The final hidden state of LSTM model, denoted as *E*_*ts*_, serves as the embedding of the time-series data with default 64 dimensions. Concurrently, *X*_*notes*_ represents clinical notes with a dimension of 512, complying with the maximum input size requirement of BioMedBERT. As illustrated in Figure 2, a necessity arises to partition the clinical notes into several subsequences to meet the specified requirement. The embedding matrix *E*_*notes*_ ∈ *R*^516×789^ is elucidated, where 512 corresponds to the dimensionality of the embedding for each token, encapsulating contextual relationships of the word within a 512-dimensional space. The value 768 designates the hidden size in the transformer model, representing the broader contextual understanding of the input sequence which determined by the transformer’s self-attention mechanism.

The calculation of *Ē*_*notes*_ involves two steps: initially, the average of the embeddings of all the subsequences from the same patient is computed, followed by taking the mean in the 512 dimensions, representing the average contribution of each dimension across all embeddings, and providing a condensed representation of the overall information captured by the embedding. Consequently, *E*_*notes*_ is a vector with 768 dimensions. The symbol ⊕ denotes the concatenate operation, wherein the embeddings from LSTM and BioMedBERT are combined to form the final embedding *E*_11_ for predictive tasks. As depicted in Figure 2, the ultimate embedding undergoes processing through a fully connected layer and a Softmax layer to derive predictions for the designated task.

## 3 Results

### 3.1 Data processing

In this study, we utilized the complete MIMIC-IV database^48^, which includes approximately 73,181 ICU stays across 50,920 critical care patients. We curated time series data from this resource inspired by prior work^41,49,50^. Figure 3 summarizes the data extraction and processing steps.

**Figure 3:**
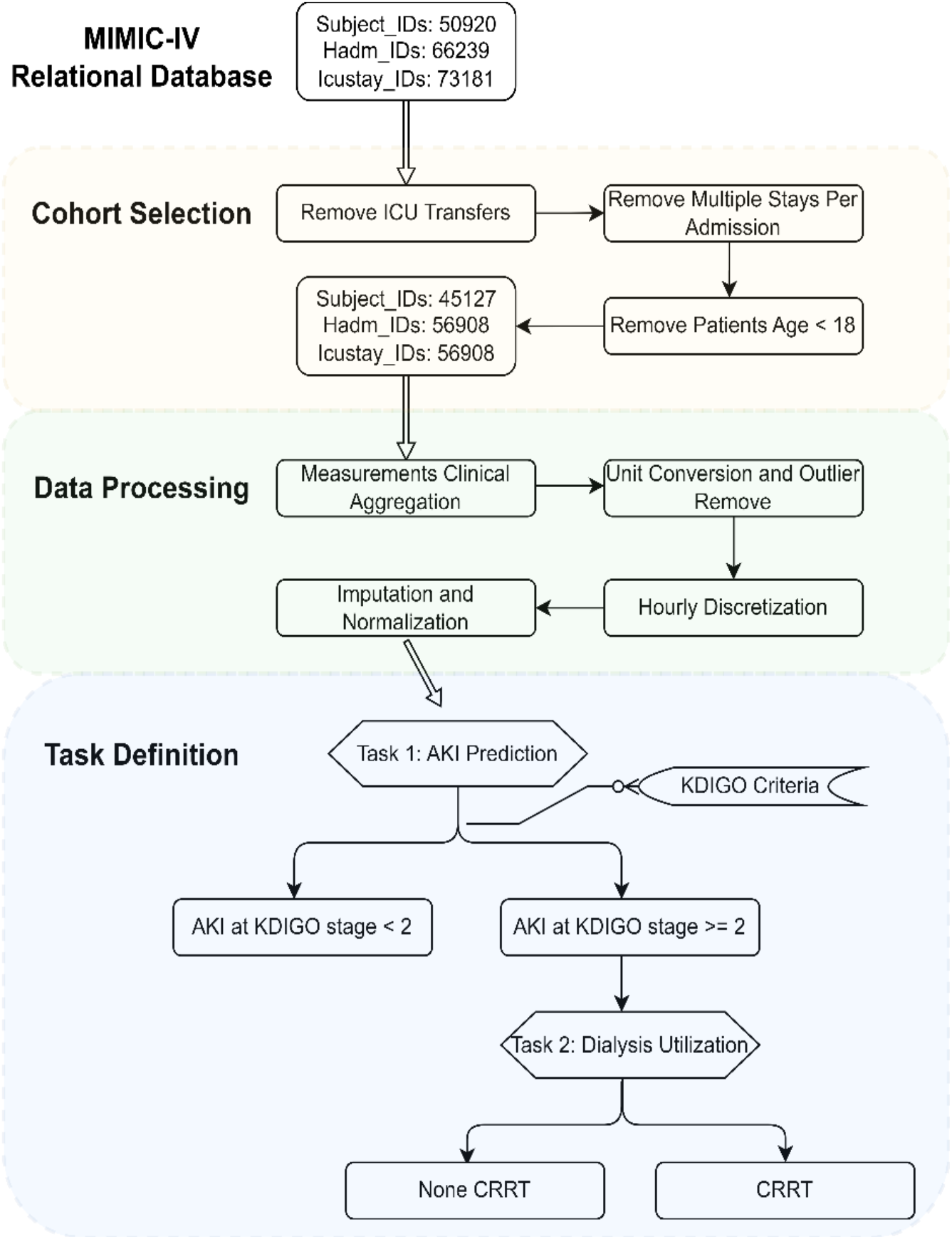
MIMIC-IV Data Processing Diagram: First, a cohort is established by applying predefined selection criteria. Next, the data undergoes manipulation involving the aggregation of measurements, unit conversion, outlier filtration, discretization, imputation, and normalization. Finally, data extraction is conducted in alignment with our task definition.

To select our cohort as illustrated in Figure 3, we extracted pertinent data from the raw MIMIC-IV tables and organizing them by patient as the initial step. Exclusion criteria are applied to admissions and ICU stays, first excluding hospital admissions with multiple ICU stays or transfers between different ICU units to mitigate outcome ambiguity associated with hospital admissions. Additionally, we excluded ICU stays involving patients under 18 due to significant physiological differences between adults and pediatric patients. The resulting root cohort comprises 45,127 unique patients, with a total of 56,908 ICU stays and over 300 million clinical events.

We next moved into the data processing phase, where clinical aggregation is initially performed. Each measurement in the MIMIC-IV database is associated with a unique ItemID, but multiple ItemIDs may match to semantically equivalent clinical features because of the different version of EHR or the same measurement at various places, e.g., admission and ICU. For example, “Weight” may be recorded under 224639, 226512, or 226531. Then a clinical taxonomy^49^ is employed to group semantically equivalent ItemIDs into more robust “clinical aggregate” features, thereby reducing data missingness and duplicate measures. Subsequently, unit conversion and outlier detection are executed to address data unit inconsistencies and handle outliers. Measurements are standardized into consistent units, with clinical experts’ input used to define valid clinical measure ranges and thresholds for detecting unusable outliers^41^. This comprehensive data curation ensures the integrity and quality of the dataset for subsequent analyses.

After the data cleaning phase, we generated time-series data and imputed missing values. In the MIMIC-IV raw data, fine-grained timestamps are provided for each laboratory measurement and recorded vital sign. However, the frequency of most measurements is notably sparse, with certain variables having infrequent occurrences. To create a more interpretable and machine-learning-friendly representation, we employed an aggregation strategy for the observations within each ICU stay’s time-series.

Specifically, we discretized the data into hourly buckets, facilitating a denser representation conducive to modern machine learning methods designed for time-series analyses that expect discretized time representations. In cases where multiple measurements of the same variable are within a single hour, we selected the measurement that is nearest to the discrete integer time point. Conversely, if no measurement occurred within an hour, we resorted to imputation by selecting the value from the previous hour. In the last step, we performed normalization, ensuring that the time-series data is scaled for seamless integration into the subsequent modeling stages.

For the clinical notes, while the patients typically possess a myriad of diverse notes, the inherent constraints of BERT-based models necessitate adherence to a fixed maximum input sequence length. To address this, we systematically divided notes into subsequences, each containing up to 300 words. It is pertinent to note that BERT processes input using sub-word units (WordPieces) rather than entire words. Given BERT’s restriction to a maximum sequence of 512 sub-word unit tokens, equivalent to approximately 300 words, we aligned our subsequence length accordingly. In addition, we kept the relevant information of the clinical notes (described in 2.2 Methodological framework section) and converted words to lowercase, removing line breaks and carriage returns, de-identifying brackets, and eliminating special characters such as “==“, “--” ^45^.

### 3.2 Generation of training and testing data

Guided by the task definition in the Figure 3 and description in 2.2 Methodological framework section, we generated training and testing datasets tailored for AKI and CRRT prediction. Given that patients with time-series data and with clinical notes do not exhibit a one-to-one correspondence, we selected patients who possess both types of information. Subsequently, we randomly allocated approximately 80% of the events as training data and the remaining 20% as testing data for both AKI and CRRT prediction. The resultant dataset comprised 38,400 records for AKI training data, of which 14,168 are positive instances. For AKI testing data, we had 10,586 records, with 2,103 of them being positive instances. Similarly, the CRRT training data encompassed 18,697 records, with 764 positive instances. The CRRT testing data comprised 4,473 records, with 52 positive instances. It is imperative to highlight the inherent challenge of class imbalance in CRRT prediction. The marked disproportion in the number of positive instances poses an additional layer of complexity to the predictive modeling task.

### 3.3 Performance comparison

Next, we evaluated and compared the performance of corresponding unimodal models on the same tasks across the two distinct data types with the goal of identifying the most effective models within their respective domains to serve as baselines for comparison with the performance of the multimodal model we have proposed.

#### 3.3.1 Time-series unimodal models’ comparison

The results presented in Table 1 provide a comparative analysis of unimodal models utilizing time-series data. Notably, LSTM consistently outperforms the other two models across both tasks, particularly when evaluating metrics such as AUPRC, precision, and recall.

#### 3.3.2 Clinical notes unimodal models’ comparison

Table 2 illustrate the performance comparison of unimodal models utilizing clinical notes data. Although the ClinicalBERT model exhibits slightly superior recall, considering numerous factors, especially the AUPRC, which is indicative of performance in imbalanced data scenarios, we conclude that BioMedBERT is the more suitable model for this comparison.

**Table 2.** BioMedBERT and ClinicalBERT performance comparison for (a) AKI (b) CRRT prediction.

| Model | Accuracy | Precision | Recall | AUROC | AUPRC |
| --- | --- | --- | --- | --- | --- |
| BioMedBERT | **0.727** | **0.376** | 0.542 | **0.742** | **0.420** |
| ClinicalBERT | 0.683 | 0.334 | **0.578** | 0.705 | 0.357 |
| (a) |  |  |  |  |  |
| Model | Accuracy | Precision | Recall | AUROC | AUPRC |
| BioMedBERT | **0.989** | **0.556** | **0.294** | **0.970** | **0.411** |
| ClinicalBERT | 0.987 | 0.353 | 0.118 | 0.924 | 0.219 |
| (b) |  |  |  |  |  |

#### 3.3.3 Performance of multimodal model

Next, we compared the performance of our multimodal model with that of the top-performing unimodal model. The results as detailed in Table 3 and Figure 4 consistently highlights the superior efficacy of the multimodal model with p-value 3.7x10^−3^ (AKI) and 2.68x10^−5^ (CRRT) of McNemar’s test comparing with LSTM. Analysis of precision and recall in CRRT predictions (Table 3(b)) indicates that our multimodal model identifies over 25% more patients who require dialysis than the unimodal model (LSTM). This translates to an increase in prediction accuracy for CRRT of approximately 25%, a significant improvement with the potential to reduce hospital complications and improve patient outcomes.

**Table 3.** Multimodal and unimodal model performance comparison for (a) AKI (b) CRRT prediction.

| Model | Accuracy | Precision | Recall | AUROC | AUPRC |
| --- | --- | --- | --- | --- | --- |
| Multi-Modal | **0.860** | **0.642** | **0.693** | **0.888** | **0.727** |
| LSTM | 0.854 | 0.628 | 0.673 | 0.873 | 0.699 |
| BioMedBERT | 0.727 | 0.376 | 0.542 | 0.742 | 0.420 |
| (a) |  |  |  |  |  |
| Model | Accuracy | Precision | Recall | AUROC | AUPRC |
| Multi-Modal | **0.997** | **0.895** | **0.829** | **0.997** | **0.840** |
| LSTM | 0.992 | 0.595 | 0.537 | 0.985 | 0.603 |
| BioMedBERT | 0.989 | 0.556 | 0.294 | 0.970 | 0.411 |
| (b) |  |  |  |  |  |

**Figure 4.**
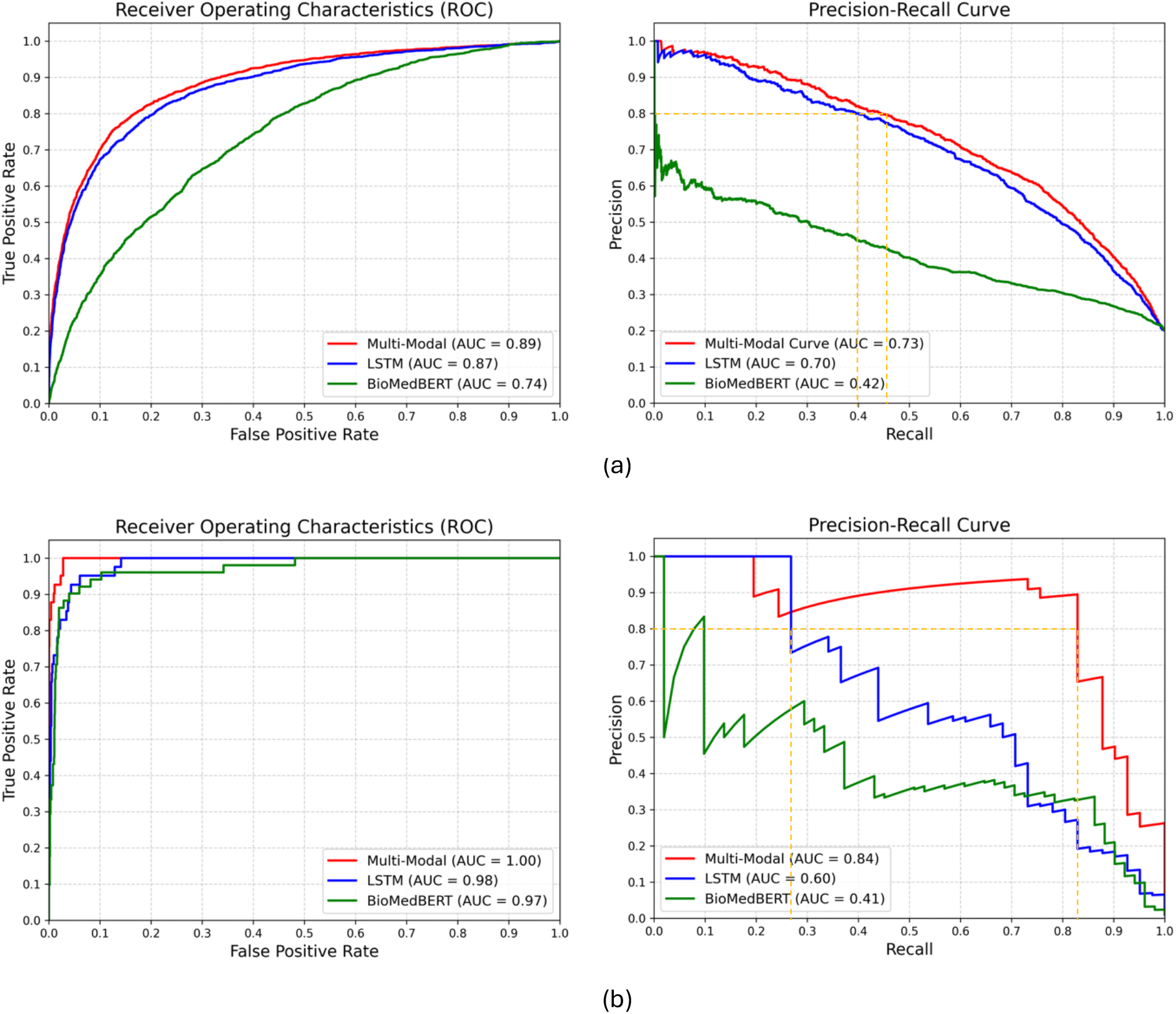
Multimodal and unimodal model performance comparison for (a) AKI (b) CRRT prediction.

Moreover, the AUPRC (Figure 4(b)) for our multimodal model shows an improvement of more than 20% over the unimodal model (LSTM), suggesting that the multimodal model can effectively minimize unnecessary treatments or interventions while accurately identifying critical cases. This finding is critical in the context of CRRT data because it is highly imbalanced and provides limited input for unimodal models. The strength of our multimodal approach is its ability to leverage complementary insights from different data sources, leading to a more comprehensive understanding of underlying patient data patterns.

For clinical tasks, where false positives are of paramount concern, a precision threshold of 80% is set to minimize the risk of alarm fatigue. Our results, specifically AUPRC plots in Figure 4(b), show that at the level of precision, our multimodal model achieves almost 50% higher recall than the unimodal model (yellow dotted line). This strategic evaluation approach ensures the minimization of false positives while optimizing recall, contributing to the clinical utility and reliability of the unimodal model in real-world applications.

### 3.4 Elucidating Feature Importance in AKI and CRRT Prediction Using SHAP Values

Building upon the enhanced performance of our multimodal model, we prioritized interpretability, which is essential for clinical adoption. Leveraging concepts from Integrated Gradients and SmoothGrad^51,52^, the integration of SHapley Additive exPlanation (SHAP) values with the expected gradients algorithm allowed us to dissect the model’s predictive mechanics at various time points, providing actionable insights into the features most contributory to the risk of AKI and CRRT requirement.

Analysis of the top SHAP values across six time points reveals that the model assigns varying degrees of importance to different clinical features in the prediction of AKI (Figure 5(a)). These features include physiological parameters, laboratory results, and patient vitals, which are known to be associated with kidney function and AKI risk. At the first time-point; we identified urine output, oxygen saturation, and diastolic blood pressure as the most influential features. Urine output is a direct measure of kidney function, with deviations from normal ranges indicating potential renal compromise. Oxygen saturation reflects systemic oxygen delivery, where hypoxia may contribute to renal ischemia^53^. Diastolic blood pressure is indicative of renal perfusion, with both hypotension and hypertension potentially leading to AKI. As the patients’ clinical course progresses at the second timepoint, urine output remains a predominant feature, emphasizing its critical role in ongoing AKI risk assessment. Oxygen saturation and diastolic blood pressure continue to be significant, alongside red blood cell count, which may indicate changes in blood volume or hematologic conditions affecting kidney function. By the third timepoint, additionally, red blood cell count, and weight emerge as key indicators, with weight potentially reflecting fluid balance, a crucial factor in AKI development^54^. Respiratory rate and neutrophils also rank highly, the former potentially signifying respiratory compensation for metabolic acidosis, and the latter suggesting an inflammatory or infectious process that could impact kidney health. At the fourth timepoint, urine output, red blood cell count, and oxygen saturation continue to dominate the model’s attention, sustaining their roles as essential markers for AKI prediction. Notably, albumin and lactate levels come into focus, where hypoalbuminemia can be associated with chronic illness impacting the kidneys^55^, and elevated lactate may indicate tissue hypoxia and metabolic distress, often preceding renal injury^56,57^. By the 5^th^ and 6^th^ time-points, the significance of urine output in AKI prediction is highlighted as it remains as the feature with the highest SHAP value.

**Figure 5:**
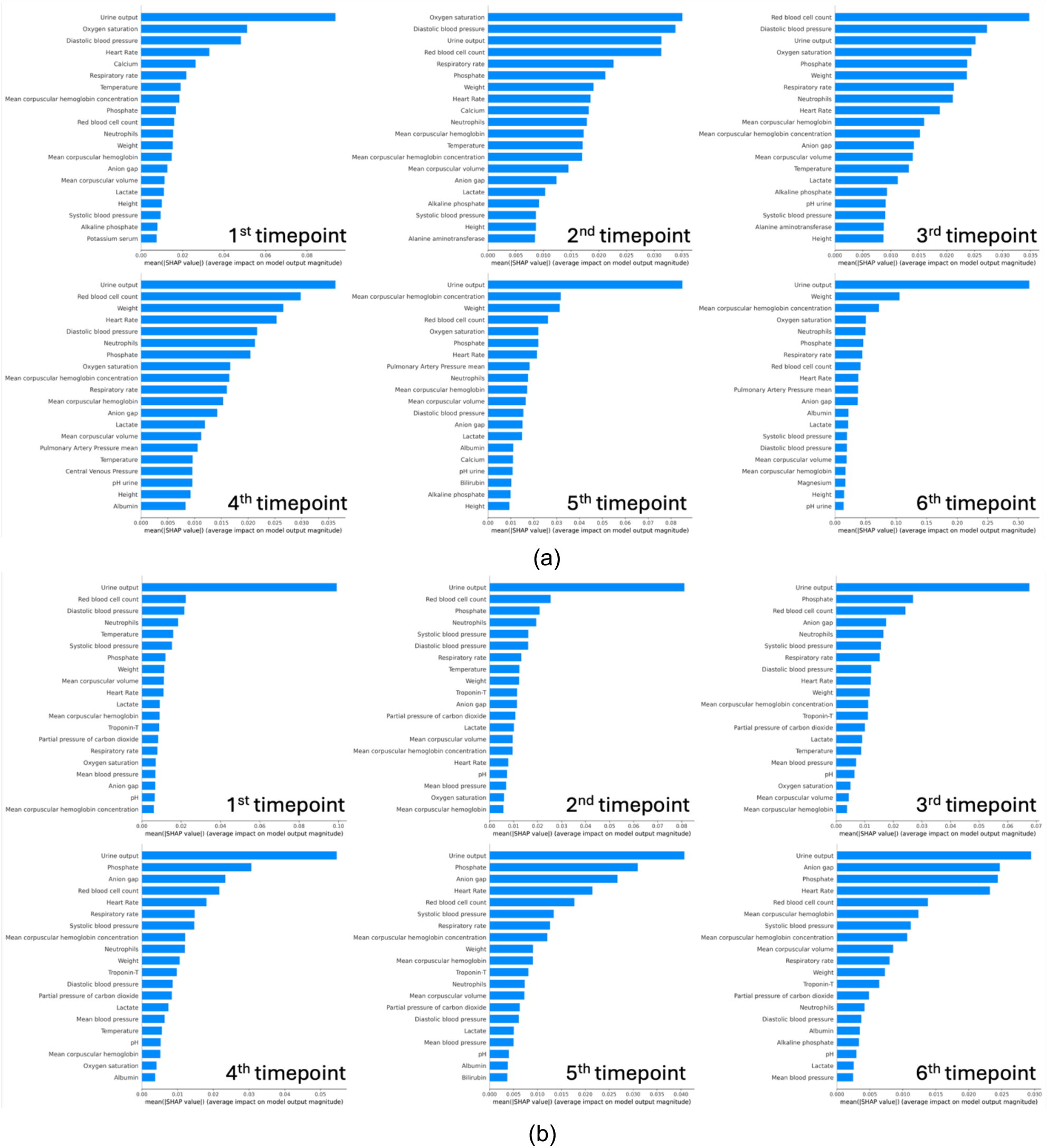
(a) AKI (b) CRRT prediction SHAP values across timepoints

The consistent prominence of urine output across all timepoints stresses its essential role in assessing renal function and in AKI staging criteria. The presence of vital signs such as blood pressure and oxygen saturation across multiple timepoints highlights the systemic nature of AKI, where renal health is closely tied to overall hemodynamic stability of the patient. Laboratory parameters such as red blood cell count and albumin levels provide insight into the patient’s volume status and nutritional state, which are integral to kidney function. These trends align with the known pathophysiology of AKI, where both acute changes and underlying chronic conditions converge to impact renal health.

Similarly for our predictive model for CRRT requirement, we observed the urine output as the most important and consistent predictor across all time points (Figure 5(b)). The anion gap is a marker for metabolic acidosis, a common occurrence in renal failure and an indicator for CRRT^58^. Beginning from the second time point, we begin observing anion gap as an important feature and its feature importance rank increases with time. Elevated phosphate levels can indicate renal dysfunction, as kidneys play a central role in phosphate regulation. Our results show phosphate levels are among the most important and persistent features that predict CRRT need in our patient cohort. By the fourth timepoint, the model highlights the anion gap and respiratory rate, the latter of which may signify respiratory compensation for metabolic acidosis—a condition often necessitating CRRT. In the final timepoint analysis, urine output, alongside anion gap and phosphate, remains a principal feature. Our analyses reveal a clear trend: certain features, particularly urine output, the anion gap, and phosphate, are consistently prominent across all timepoints in predicting the need for CRRT. This indicates a strong association of these features with the underlying pathophysiological processes leading to requirement for CRRT, such as fluid overload, electrolyte imbalances, and acid-base disturbances.

Collectively, the interpretative power of SHAP values in our models not only validates the clinical relevance of the identified features but also enhances the model’s utility by offering clinicians a detailed profile of AKI features and CRRT risk factors as they evolve over time. Recognizing these features’ significance can assist clinicians in early identification of patients who may benefit from CRRT, thereby potentially improving outcomes in critically ill patients, through timely and targeted interventions.

### 3.5 Clinical notes analysis using BioMedBERT embeddings

The incorporation of BioMedBERT allowed our multimodal model to analyze unstructured clinical notes from the MIMIC-IV database for the prediction of AKI and CRRT. Through this analysis, we identified key terms (tokens) that BioMedBERT attributed with high importance scores, potentially revealing clinical narratives associated with these conditions. Here it is important to remember that we only included notes recorded at the time of patient admission and excluded discharge notes to focus on data relevant to the initial phase of hospitalization.

Our results showed that for AKI prediction, the term ‘hemodialysis’ appeared with the highest importance score and similarly, ‘dialysis’ was captured as significant, which may initially seem inconsistent with our focus on the initial phase of hospitalization. However, these terms likely represent significant risk factors in the patient’s past medical history, which are crucial for predicting the development of AKI. The presence of these tokens shows our model’s ability to identify patients with pre-existing conditions that elevate the risk of AKI, even if the actual treatment or condition is not an immediate concern at the point of hospital admission. We also observed ‘hd’ (hemodialysis) and ‘laparotomy’ (which is a surgical procedure involving a large incision through the abdominal wall) in the token list, suggesting possible postoperative kidney stress, which is a recognized risk factor for AKI development^59,60^. Moreover, the presence of terms such as ‘ckd’ (chronic kidney disease), ‘cirrhosis,’ and ‘ascites’ indicates underlying chronic conditions that can predispose patients to AKI. Additionally, tokens like ‘jaundice,’ and ‘ed’ (emergency department) highlight symptoms and care settings that may be relevant to the patient’s acute or chronic health challenges.

For the predictions related to CRRT, the token ‘endotracheal’ is associated with the highest importance score. It is relevant as it indicates airway management typically required for critically ill patients, who may also be at an elevated risk for CRRT. The presence of ‘obese’ and ‘intubation’ as important tokens correlates with known risk factors for AKI, which can lead to CRRT. Obesity is associated with several comorbidities that predispose individuals to kidney stress^61^, and intubation is often part of the management of severely ill patients who are at an increased risk for renal complications.

Overall, our multimodal model’s ability to identify significant tokens from clinical admission notes offers a promising avenue for enhancing patient risk assessment during the critical initial phase of hospitalization. The detection of terms related to prior renal health problems provides a deeper insight into the patient’s condition, facilitating a proactive approach to AKI and CRRT management. This can be particularly beneficial for tailoring both monitoring and treatment strategies, potentially reducing the progression to severe renal events.

## 4. Conclusions

In this study, we developed a longitudinal, multimodal model that capitalizes on both unstructured clinical notes and structured time-series measurements derived from EHRs for the early AKI prediction and the prospective identification of the necessity for CRRT. Our model demonstrates exceptional predictive accuracy, offering valuable insights into AKI and CRRT needs at least 12 hours ahead of clinical manifestation. The achieved AUROC and AUPRC metrics further underscore the robustness of our system. The incorporation of interpretability measures, including SHAP values and the expected gradients algorithm, ensures transparency and positions our model as a reliable candidate for clinical deployment. This research holds promise in optimizing AKI management, contributing to improved patient outcomes, and marks a significant stride toward enhancing healthcare practices.

## Data Availability

All data produced are available online at MIMIC-IV

## CRediT authorship contribution statement

**Yukun Tan:** Conceptualization, Data curation, Formal analysis, Investigation, Methodology, Software, Visualization, Validation, Writing - original draft, Writing - review & editing. **Merve Dede:** Conceptualization, Investigation, Formal analysis, Validation, Writing - original draft, Writing - review & editing. **Vakul Mohanty:** Conceptualization, Visualization. **Jinzhuang Dou:** Conceptualization, Software. **Holly Hill:** Conceptualization, Writing - review & editing. **Elmer Bernstam:** Supervision, Validation, Writing - review & editing. **Ken Chen:** Conceptualization, Supervision, Project administration, Funding acquisition, Writing – review & editing.

